# The CorNeA Registry: Design, Governance, and Early Multicenter Characteristics of an International Registry for Corneal Neurotization in Neurotrophic Keratopathy

**DOI:** 10.64898/2026.04.22.26351277

**Authors:** Pareena Sharma, Kareem Wali, Jordan Crabtree, Khoa Tran, Sara Williams, Charline S. Boente, Konstantin Feinberg, Asim Ali, Gregory H. Borschel

**Affiliations:** Indiana University School of Medicine and Riley Children’s Hospital, Division of Plastic Surgery, Indianapolis, Indiana, USA; Indiana University School of Medicine and Riley Children’s Hospital, Department of Ophthalmology, Indianapolis, Indiana, USA; University of Toronto and the Hospital for Sick Children, Department of Ophthalmology, Toronto, Canada

**Keywords:** corneal neurotization, neurotrophic keratopathy, neurotrophic keratitis, corneal sensation, corneal reinnervation, corneal ulcer, nerve graft

## Abstract

**Background:** Neurotrophic keratopathy (NK) is a rare degenerative corneal disease caused by impaired trigeminal innervation, resulting in reduced corneal sensation, impaired epithelial healing, ulceration, and risk of perforation or vision loss. Corneal innervation is essential for protective reflexes, epithelial maintenance, and ocular surface homeostasis. Conventional medical therapies may promote epithelial healing but do not directly restore corneal innervation. Corneal neurotization (CN) has emerged as a surgical strategy in which healthy donor sensory axons are transferred to denervated corneas to provide innervation. Multiple procedural variations now exist, including differences in donor nerve selection, graft use, and methods of limbal nerve insertion. A broad variety of NK etiologies is also being treated, including congenital, infectious, tumor, or other causes. However, published evidence remains limited by small case series, heterogeneous surgical methods, short follow-up periods, and inconsistent outcome reporting.

**Objective:** To address the need for standardized long-term outcome data in CN, we established the Corneal Neurotization Assessment (CorNeA) Registry, an international multicenter observational registry designed to evaluate patients undergoing CN.

**Methods:** The CorNeA Registry captures demographic characteristics, disease etiology, surgical technique, and longitudinal ocular outcomes in patients with NK treated with CN. Data are recorded in REDCap and include both retrospective and prospective patient enrollment across participating centers. Patients are followed longitudinally after surgery without a predefined endpoint to permit long-term assessment of corneal sensation recovery, ulcer recurrence, and visual outcomes. At the time of reporting, the registry includes 58 patients from multiple international centers, with active expansion ongoing.

**Conclusion:** Because NK is rare and CN remains an evolving surgical field, long-term comparative data are lacking. The CorNeA Registry provides the first international platform to characterize patient selection, procedural variation, and long-term outcomes after CN, with the goal of informing future surgical decision-making and outcome standardization.

## Introduction

Neurotrophic keratopathy (NK) is a rare condition affecting 5 in 10,000 people in which innervation to the cornea is diminished or absent, causing deficits in corneal sensation, corneal wound healing and maintenance^1–4^. The cornea receives its innervation from the ophthalmic branch of the trigeminal nerve, and corneal sensation is vital for protective eye reflexes like blinking and tearing. Without corneal innervation, nerve-derived neurotrophic factors that maintain corneal homeostasis through epithelial renewal and healing are altered^5^. Lack of corneal sensation causes a degenerative state in which corneal thinning and ulcers may result in scarring, vision loss, and perforation. Due to its chronic nature, NK is notoriously difficult to treat and has a large impact on quality of life^1^.

Currently, there is no gold standard treatment for NK. Traditional treatments include protective lenses, lubricating drops, biologic agents, punctal plugs, amniotic membrane transplantation, and tarsorrhaphy. While these can help delay disease progression, they do not address the underlying issue of corneal denervation^1^. Corneal neurotization (CN) is a novel procedure in which healthy donor nerve fibers are surgically transferred to a denervated cornea to achieve innervation, seeking to directly address the underlying cause of NK. CN promotes long-term healing of persistent epithelial defects, provides corneal sensation in many cases, and may lead to secondary improvements in visual function, while also enabling subsequent interventions such as corneal transplantation^8–13^.

In 2009, Terzis reported direct corneal neurotization (DCN) by transferring contralateral supraorbital and supratrochlear nerves into the affected cornea’s perilimbal area^17^. Many groups have reported different techniques^8–13,18^, and variations in donor sensory nerve source, use of nerve grafts, nerve coaptation method, and nerve insertion technique into the limbal area. ^19,20^.

Currently, there is no database to evaluate outcomes of corneal neurotization surgery. Studies have been limited to small case series typically reporting single surgeon’s results with a wide variation in procedure and patient characteristics. Additionally, long-term effectiveness of CN is rarely reported. A recent meta-analysis of CN outcomes by Swanson et al. noted a median follow-up period of only 13 months^21^.

CN presents unusual challenges for conventional single-center studies because NK itself is rare, etiologies are heterogeneous, and both pediatric and adult populations are affected. In addition, important clinical decisions often occur outside conventional surgical variables, including whether intervention should occur before recurrent ulceration, after failure of medical therapy, or in sequenced with corneal transplantation. These questions are particularly consequential in pediatric patients, where visual development, amblyopia risk, and sensory cortical plasticity may influence both timing and outcome.

The absence of standardized multicenter data has limited the ability to determine whether timing of intervention, donor nerve choice, graft strategy, or surgical approach influence long-term corneal recovery. To address these concerns, we established the Corneal Neurotization Assessment (CorNeA) Registry, an international multicenter observational registry designed to capture longitudinal clinical data in patients undergoing CN. By standardizing data collection across institutions and surgical approaches, the registry is intended to characterize procedural variation, define longitudinal outcomes, and support future comparative analyses across the evolving spectrum of CN techniques.

## Methods

### Registry Design

The CorNeA Registry is an international, multicenter, longitudinal observational registry of pediatric and adult patients with neurotrophic keratopathy (NK) undergoing corneal neurotization (CN) (Figure 1). The registry was established to address the absence of standardized long-term outcome data in CN, a rapidly evolving surgical field characterized by procedural heterogeneity and limited comparative evidence. Data are collected prospectively in patients enrolled before surgery and retrospectively in patients who underwent CN before registry activation at a participating site. Longitudinal follow-up continues for a minimum of 3 years after surgery, with no predefined endpoint for continued data collection, permitting assessment of delayed sensory recovery, epithelial stability, and long-term ocular outcomes.

**Figure 1.**
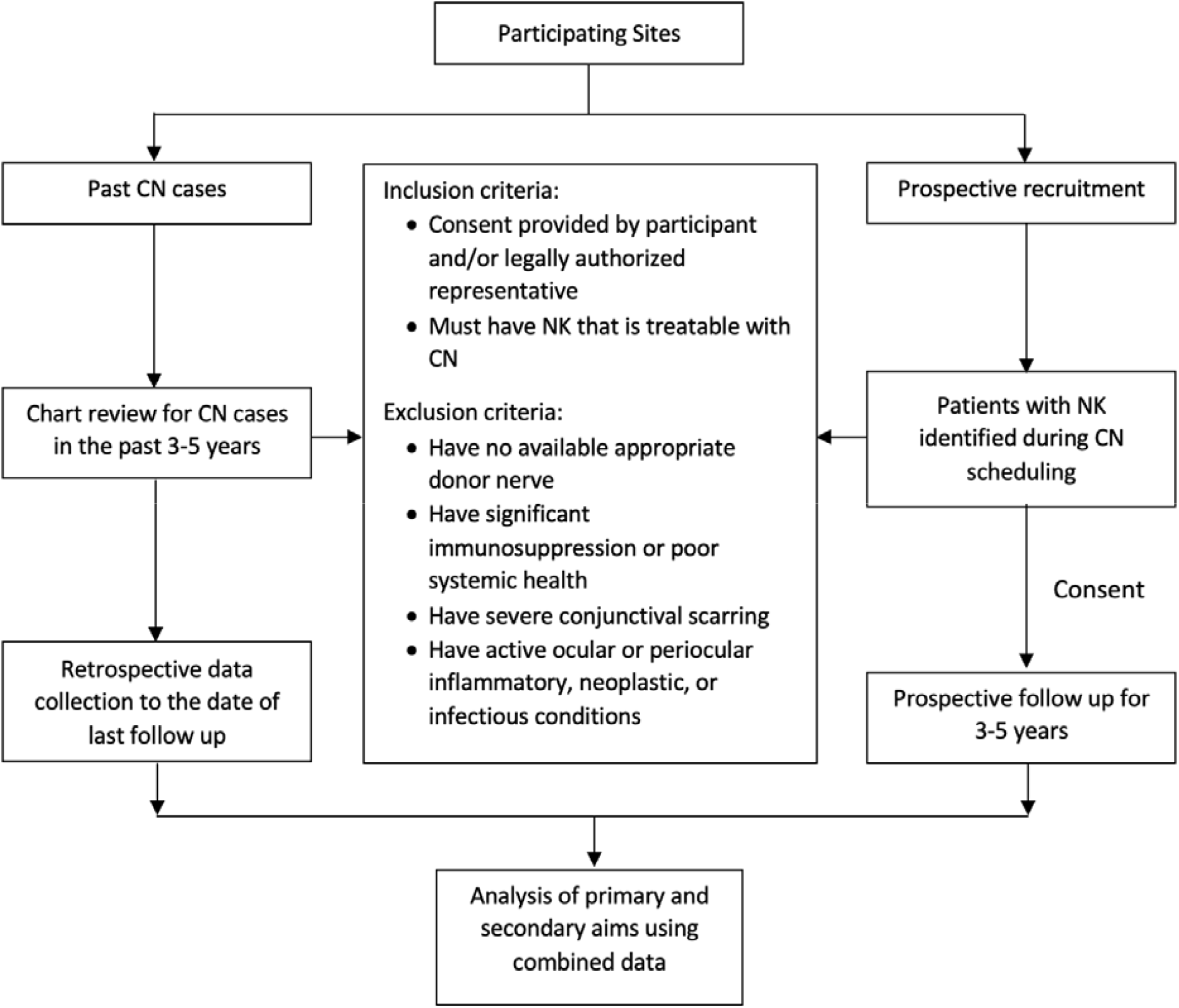
CorNeA Registry study design. Flowchart illustrating the dual-arm study design of the CorNeA Registry across participating sites. The retrospective arm identifies past CN cases through chart review spanning 3–5 years prior to enrollment, with retrospective data collected through the date of last follow-up. The prospective arm recruits patients with neurotrophic keratopathy (NK) identified during CN scheduling, who provide informed consent and undergo prospective follow-up for 3–5 years. Eligibility for both arms is governed by uniform inclusion criteria (consent provision and presence of NK treatable with CN) and exclusion criteria (absence of a suitable donor nerve, significant immunosuppression or poor systemic health, severe conjunctival scarring, or active ocular/periocular inflammatory, neoplastic, or infectious conditions). Data from both arms are combined for analysis of primary and secondary study aims.

### Participating Sites and Oversight

Indiana University School of Medicine serves as the coordinating center for the CorNeA Registry. Registry governance is structured through three coordinating bodies: a Steering Committee, a Scientific Advisory Committee, and an Operations Committee (Figure 2). The Steering Committee provides overall scientific oversight, approves participating sites and research proposals, and guides publication priorities. The Scientific Advisory Committee reviews proposed projects and identifies emerging scientific questions suitable for registry-based investigation. The Operations Committee, which includes site investigators, coordinators, and biostatistical support from Regenstrief Institute, oversees data harmonization, quality assurance, and operational coordination across participating centers.

**Figure 2.**
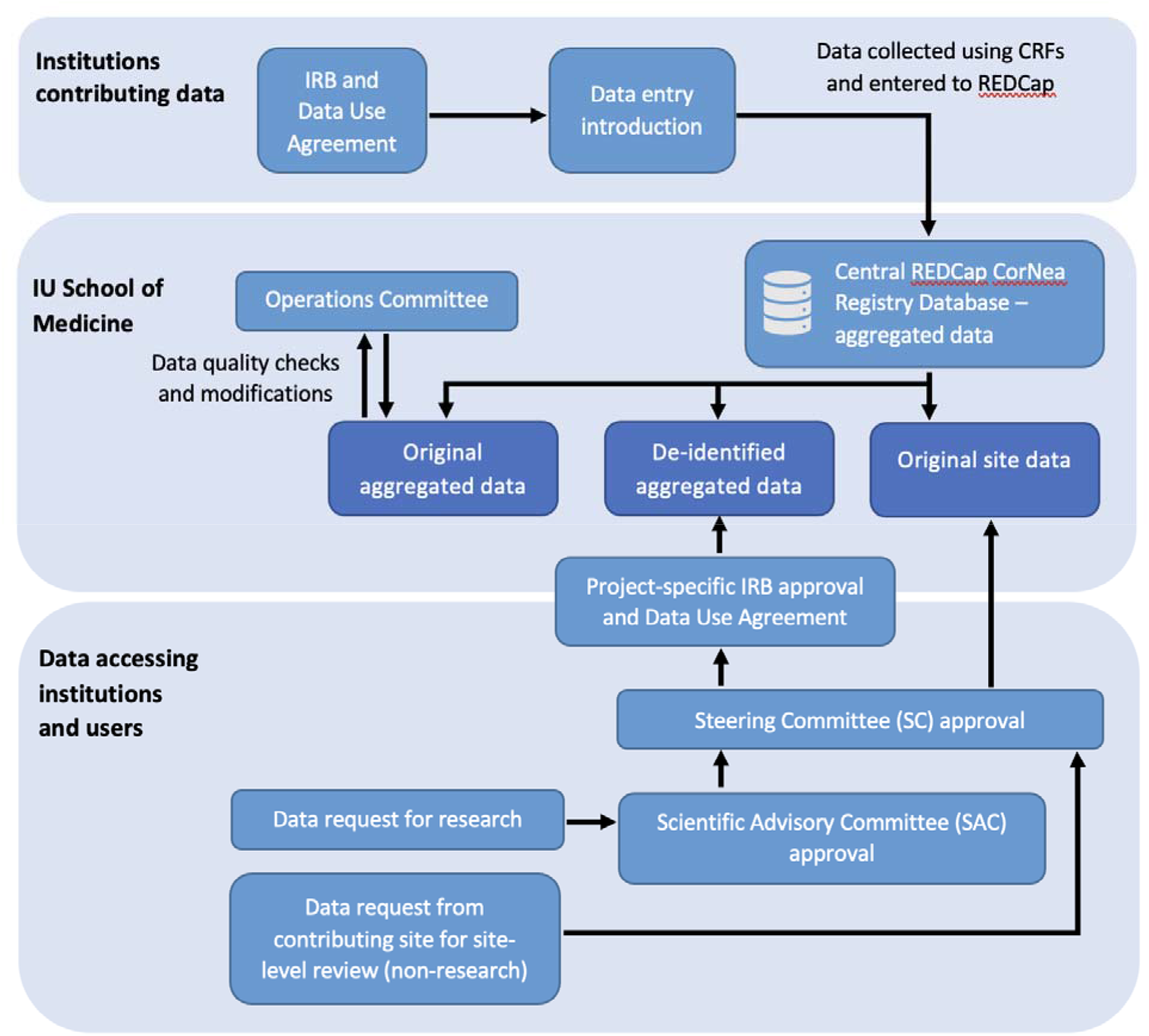
CorNeA Registry regulatory framework and data access structure. Schematic illustrating the three-tier governance model governing data contribution and access within the CorNeA Registry. Contributing institutions execute an IRB approval and Data Use Agreement (DUA) with IU School of Medicine, after which data are collected via case report forms (CRFs) and entered into the central REDCap CorNeA Registry database. The IU School of Medicine Operations Committee oversees ongoing data quality checks and modifications to the aggregated dataset. Institutions and investigators seeking data access, participation as a contributing site is not required for data access, submit a project-specific Data Request, which undergoes sequential review by the Scientific Advisory Committee (SAC) and Steering Committee (SC). Upon approval, a project-specific IRB approval and DUA are executed, and access is granted to the appropriate data tier: original aggregated data, de-identified aggregated data, or original site data. Contributing sites may additionally request site-level data review for non-research purposes.

Participating sites are recruited through investigator invitation and referral from surgeons performing CN internationally. Before activation, each site obtains local regulatory approval, executes a data use agreement, and completes REDCap training to standardize data interpretation and entry procedures. At the time of reporting, 5 additional sites were actively completing regulatory onboarding, and 47 sites across 20 countries had expressed formal interest in future participation, reflecting continued international expansion (Figure 3).

**Figure 3.**
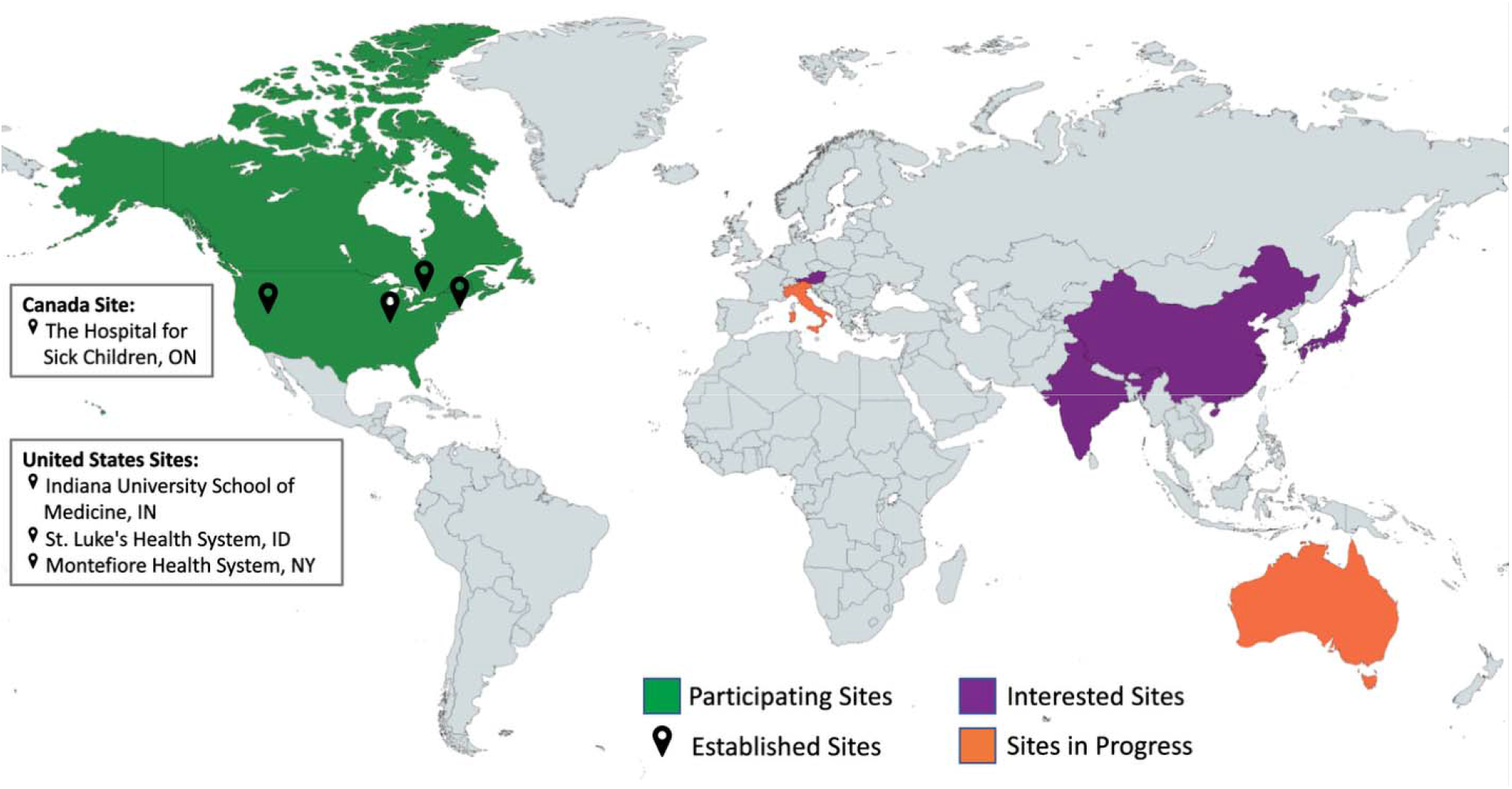
International Site Map of the CorNeA Registry. Registry sites are categorized into three groups: fully enrolled participating sites (green; United States, Canada), sites currently undergoing enrollment (orange; Italy, Australia), and sites that have expressed formal interest in joining (purple; Austria, India, China, Japan). Established site locations within participating countries are indicated by pins (The Hospital for Sick Children, Toronto, ON; Indiana University School of Medicine, Indianapolis, IN; St. Luke’s Health System, Boise, ID; Montefiore Health System, New York, NY).

### Patient Population and Eligibility

Eligible participants include pediatric and adult patients diagnosed with NK who are considered candidates for CN by the site investigator. Enrollment may occur prospectively before surgery or retrospectively through chart review after local institutional review board approval and execution of a data use agreement.

Prospective participants provide written informed consent from the patient or legally authorized representative according to local regulatory requirements. Retrospective participants are identified through site-based chart review, with historical and longitudinal clinical data abstracted from the medical record. Patient care follows local standards and is not altered by registry participation.

Participants are eligible if they meet all the following criteria:

1. Diagnosis of neurotrophic keratopathy
2. NK considered amenable to treatment with corneal neurotization
3. Provision of informed consent by the participant or legally authorized representative in accordance with local requirements
4. Ability to participate in follow-up assessment

Participants are excluded if any of the following are present:

1. Absence of a suitable donor sensory nerve or anatomic limitation precluding CN
2. Significant immunosuppression or poor systemic health
3. Severe conjunctival scarring
4. Active ocular or periocular inflammatory, infectious, or neoplastic disease

Participants may undergo any CN technique, including direct or indirect approaches, regardless of donor sensory nerve source, graft type, coaptation method, or limbal insertion strategy, permitting capture of contemporary procedural diversity across centers. Participants may withdraw from the registry at any time, with previously collected data retained unless removal is specifically requested.

### Outcomes

Primary registry outcomes include postoperative recovery of corneal sensation and the presence or recurrence of corneal ulceration. Corneal sensation is assessed using Cochet-Bonnet aesthesiometer when available, with cotton-wisp testing recorded when quantitative aesthesiometry is not feasible. Corneal epithelial status is documented at each follow-up visit, including fluorescein findings when available, together with interval ulcer recurrence since the preceding visit.

Secondary variables include demographic characteristics, NK etiology, ocular comorbidities, operative details, and postoperative recovery measures. Because CN outcomes may extend beyond epithelial healing alone, patient-reported outcomes are collected longitudinally using the PROMIS Global Health instrument to assess physical, mental, and social health. Registry variables were selected to support future analyses of timing of intervention, donor nerve selection, graft strategy, and durability of corneal reinnervation across evolving CN techniques and patient populations.

### Data Collection and Quality Assurance

Data are entered into a secure REDCap database using standardized electronic case report forms after site training. Each participant is assigned a global unique identifier, with identifying information retained locally at each site. Access is restricted through site-specific data access controls that permit investigators to view and edit only their own institutional data.

Automated validation within REDCap identifies incomplete entries, invalid field types, and inconsistent date ranges at the time of entry. Participating sites remain responsible for data accuracy and periodic verification, while the coordinating center performs regular cross-site data quality and accrual review. Protected health information required for longitudinal analysis, including birth and visit dates, is de-identified before aggregate data sharing to preserve patient confidentiality.

Individual sites retain ownership of their own patient data while agreeing to data sharing for approved registry-based projects. Research proposals are reviewed by the Scientific Advisory Committee and Steering Committee before aggregate data release, with project tracking used to minimize overlap and encourage multicenter collaboration.

## Statistical Analysis

Initial analyses focus on descriptive characterization of the registry cohort and examination of associations between patient characteristics, operative variables, and longitudinal clinical outcomes. Continuous variables are summarized using medians with ranges or means with standard deviations as appropriate, and categorical variables are summarized as frequencies and proportions. Time-to-event analyses, including time to recovery of corneal sensation and ulcer recurrence, are performed using Kaplan– Meier methods with log-rank testing for between-group comparisons.

To evaluate heterogeneity in postoperative recovery, group-based trajectory modeling is planned to identify distinct patterns of sensory recovery over time according to patient age, disease etiology, surgical technique, and postoperative management. As registry maturity increases, multivariable analyses will be used to explore independent associations between procedural variation and longitudinal outcomes, with particular attention to factors influencing durability of corneal reinnervation and ocular surface stability.

## Results

At the time of analysis, four sites had obtained regulatory approval and three sites had contributed patient-level data to the registry. Participating centers included university-affiliated pediatric referral hospitals and academic health systems in North America, reflecting the specialized multidisciplinary infrastructure required for corneal neurotization. Enrolling sites are geographically distributed across the Midwestern, Northeastern, and Western United States, as well as Canada. In addition, 5 sites were actively completing data use agreements, and 47 sites across 20 countries had expressed formal interest in future participation, indicating continued international expansion of the registry

The registry has already supported multiple active lines of investigation, including case reports, case series, cohort-based outcome analyses, qualitative interview studies, health economic analyses, and exploratory translational studies evaluating tear proteomic profiles in relation to clinical characteristics. These early projects illustrate the registry’s capacity to support both clinical and translational investigation across participating centers.

At the time of reporting, the registry included 58 patients undergoing corneal neurotization for neurotrophic keratopathy. Median age at enrollment was 3 years (range 4 months–23 years), reflecting the substantial pediatric representation within the cohort. Thirty-two patients (56%) were male and 26 (44%) were female. Congenital etiologies accounted for the majority of cases (36 patients, 62%), followed by brain tumor–related trigeminal injury (5 patients, 9%), ocular surgery–associated injury (4 patients, 7%), infectious causes (4 patients, 7%), trauma (2 patients, 3%), and other or unspecified etiologies (7 patients, 12%). Bilateral disease was present in 27 patients (47%), while 10 patients (17%) had isolated right-eye involvement and 21 (36%) had isolated left-eye involvement. Among 32 patients with documented amblyopia assessment, 11 (34%) were found to have amblyopia.

## Discussion

The CorNeA Registry was developed to address a central limitation in contemporary corneal neurotization research: despite increasing international adoption of CN, available evidence remains fragmented across small retrospective case series with heterogeneous patient populations, surgical methods, and outcome definitions. Existing reports differ substantially in donor nerve selection, graft use, sensory testing methodology, follow-up duration, and criteria used to define treatment success, limiting direct comparison across centers. By establishing a standardized international registry with longitudinal follow-up, the CorNeA Registry provides a framework for evaluating procedural variation and long-term outcomes within an emerging surgical field that has matured beyond proof-of-concept but remains methodologically underpowered for comparative inference.

CN presents unusual challenges for conventional clinical study because NK is rare, etiologically heterogeneous, and treated across both pediatric and adult populations. Important treatment decisions frequently extend beyond conventional operative variables and include timing of intervention relative to recurrent epithelial breakdown, medical treatment failure, amblyopia risk, and the need for corneal transplantation. These factors are difficult to evaluate within single-center cohorts but become accessible through multicenter longitudinal data collection. The predominance of congenital etiologies and early childhood enrollment within the current registry cohort further underscores the importance of pediatric representation in future analyses, particularly given the potential influence of sensory cortical plasticity and visual development on postoperative outcomes.

The registry also establishes a collaborative infrastructure capable of supporting future comparative analyses that have not previously been feasible in CN, including evaluation of donor sensory nerve selection, direct versus indirect neurotization strategies, graft choice, bilateral disease management, and timing of intervention relative to ocular surface deterioration or corneal transplantation. Because participating centers differ in both patient referral patterns and operative preference, the registry may allow emerging practice patterns to be examined under real-world conditions while preserving the diversity of contemporary surgical approaches.

Patients with NK frequently experience morbidity beyond corneal ulceration alone, including recurrent procedures, prolonged topical therapy, reduced visual function, and, in some cases, socially visible eyelid procedures such as tarsorrhaphy. Although epithelial healing is often used as a primary clinical endpoint, quality of life remains underreported in published CN studies. Incorporation of PROMIS measures into the registry therefore offers an opportunity to evaluate physical, social, and emotional outcomes alongside sensory and ocular surface recovery, expanding assessment beyond traditional ophthalmic endpoints.^28^

## Conclusion

As international participation expands, the CorNeA Registry is positioned to define standardized reporting expectations for CN and generate evidence that may ultimately inform patient selection, timing of intervention, and comparative surgical decision-making. In a field where technical innovation has advanced more rapidly than comparative clinical evidence, longitudinal registry science may provide the most practical path toward consensus.

## Data Availability

All data produced in the present work are contained in the manuscript

## Acknowledgements

Jimmie Li and Katelyn Stevens (Indiana University School of Medicine)

---

**Table 1.** Interim patient characteristics of the corneal neurotization registry (n = 58).

| Category | Subcategory | Value |
| --- | --- | --- |
| **Demographics** | Total patients | 58 |
|  | Median age at enrollment | 3 years (4 months–23 years) |
|  | Male | 32 (56%) |
|  | Female | 26 (44%) |
| **Etiology of Corneal Anesthesia** | Congenital | 36 (62%) |
|  | Brain tumor–related trigeminal injury | 5 (9%) |
|  | Ocular surgery–associated injury | 4 (7%) |
|  | Infectious | 4 (7%) |
|  | Trauma | 2 (3%) |
|  | Other/unspecified | 7 (12%) |
| **Congenital Etiology Subtypes†** | Isolated corneal anesthesia | 1 (3%) |
|  | Isolated trigeminal nerve involvement | 2 (6%) |
|  | Syndromic diagnoses | 9 (27%) |
|  | Other congenital conditions | 27 (82%) |
| **Disease Laterality** | Bilateral | 27 (47%) |
|  | Right eye only | 10 (17%) |
|  | Left eye only | 21 (36%) |
| **Amblyopia** | Patients assessed | 32 |
|  | Patients with amblyopia | 11 (34%) |

† Percentages for congenital etiology subtypes are calculated among the 36 patients with congenital etiologies. Values may exceed 100% due to overlapping diagnostic categories.

## References

1. Dua HS, Said DG, Messmer EM, et al. Neurotrophic keratopathy. Prog Retin Eye Res. 2018;66:107–131. doi:10.1016/j.preteyeres.2018.04.003

2. Sacchetti M, Lambiase A. Diagnosis and management of neurotrophic keratitis. Clin Ophthalmol. 2014;8:571–579. doi:10.2147/OPTH.S45921

3. Bonini S, Rama P, Olzi D, Lambiase A. Neurotrophic keratitis. Eye. 2003;17(8):989–995. doi:10.1038/sj.eye.6700616

4. NaPier E, Camacho M, McDevitt TF, Sweeney AR. Neurotrophic keratopathy: current challenges and future prospects. Ann Med. 2022;54(1):666–673. doi:10.1080/07853890.2022.2045035

5. Mastropasqua L, Massaro-Giordano G, Nubile M, Sacchetti M. Understanding the Pathogenesis of Neurotrophic Keratitis: The Role of Corneal Nerves. J Cell Physiol. 2017;232(4):717–724. doi:10.1002/jcp.25623

6. Bonini S, Lambiase A, Rama P, et al. Phase II Randomized, Double-Masked, Vehicle-Controlled Trial of Recombinant Human Nerve Growth Factor for Neurotrophic Keratitis. Ophthalmology. 2018;125(9):1332–1343. doi:10.1016/j.ophtha.2018.02.022

7. Pflugfelder SC, Massaro-Giordano M, Perez VL, et al. Topical Recombinant Human Nerve Growth Factor (Cenegermin) for Neurotrophic Keratopathy: A Multicenter Randomized Vehicle-Controlled Pivotal Trial. Ophthalmology. 2020;127(1):14–26. doi:10.1016/j.ophtha.2019.08.020

8. Wolkow N, Habib LA, Yoon MK, Freitag SK. Corneal Neurotization: Review of a New Surgical Approach and Its Developments. Semin Ophthalmol. 2019;34(7-8):473–487. doi:10.1080/08820538.2019.1648692

9. Dragnea DC, Krolo I, Koppen C, Faris C, Van den Bogerd B, Ní Dhubhghaill S. Corneal Neurotization-Indications, Surgical Techniques and Outcomes. J Clin Med. 2023;12(6):2214. doi:10.3390/jcm12062214

10. Elbaz U, Bains R, Zuker RM, Borschel GH, Ali A. Restoration of corneal sensation with regional nerve transfers and nerve grafts: a new approach to a difficult problem. JAMA Ophthalmol. 2014;132(11):1289–1295. doi:10.1001/jamaophthalmol.2014.2316

11. Wisely CE, Rafailov L, Cypen S, Proia AD, Boehlke CS, Leyngold IM. Clinical and Morphologic Outcomes of Minimally Invasive Direct Corneal Neurotization. Ophthal Plast Reconstr Surg. 2020;36(5):451–457. doi:10.1097/IOP.0000000000001586

12. Sweeney AR, Wang M, Weller CL, et al. Outcomes of corneal neurotisation using processed nerve allografts: a multicentre case series. Br J Ophthalmol. 2022;106(3):326–330. doi:10.1136/bjophthalmol-2020-317361

13. Leyngold I, Weller C, Leyngold M, Tabor M. Endoscopic Corneal Neurotization: Technique and Initial Experience. Ophthal Plast Reconstr Surg. 2018;34(1):82–85. doi:10.1097/IOP.0000000000001023

14. Meals RA, Nelissen RGHH. The origin and meaning of “neurotization.” J Hand Surg. 1995;20(1):144–146. doi:10.1016/S0363-5023(05)80072-3

15. Liu CY, Arteaga AC, Fung SE, Cortina MS, Leyngold IM, Aakalu VK. Corneal neurotization for neurotrophic keratopathy: Review of surgical techniques and outcomes. Ocul Surf. 2021;20:163–172. doi:10.1016/j.jtos.2021.02.010

16. Samii M. [Operative reconstruction of injured nerves]. Langenbecks Arch Chir. 1972;332:355–362. doi:10.1007/BF01282653

17. Terzis JK, Dryer MM, Bodner BI. Corneal neurotization: a novel solution to neurotrophic keratopathy. Plast Reconstr Surg. 2009;123(1):112–120. doi:10.1097/PRS.0b013e3181904d3a

18. Fogagnolo P, Giannaccare G, Bolognesi F, et al. Direct Versus Indirect Corneal Neurotization for the Treatment of Neurotrophic Keratopathy: A Multicenter Prospective Comparative Study. Am J Ophthalmol. 2020;220:203–214. doi:10.1016/j.ajo.2020.07.003

19. Bains RD, Elbaz U, Zuker RM, Ali A, Borschel GH. Corneal neurotization from the supratrochlear nerve with sural nerve grafts: a minimally invasive approach. Plast Reconstr Surg. 2015;135(2):397e–400e. doi:10.1097/PRS.0000000000000994

20. Domeshek LF, Hunter DA, Santosa K, et al. Anatomic characteristics of supraorbital and supratrochlear nerves relevant to their use in corneal neurotization. Eye. 2019;33(3):398–403. doi:10.1038/s41433-018-0222-0

21. Swanson MA, Swanson RD, Kotha VS, et al. Corneal Neurotization: A Meta-analysis of Outcomes and Patient Selection Factors. Ann Plast Surg. 2022;88(6):687–694. doi:10.1097/SAP.0000000000003117

22. Crabtree JR, Tannir S, Tran K, Boente CS, Ali A, Borschel GH. Corneal Nerve Assessment by Aesthesiometry: History, Advancements, and Future Directions. Vision. 2024;8(2):34. doi:10.3390/vision8020034

23. Safi M, Rose-Nussbaumer J. Overview of Neurotrophic Keratopathy and a Stage-Based Approach to Its Management. Eye Contact Lens. 2021;47(3):140–143. doi:10.1097/ICL.0000000000000760

24. Trinh T, Mimouni M, Santaella G, Cohen E, Chan CC. Surgical Management of the Ocular Surface in Neurotrophic Keratopathy: Amniotic Membrane, Conjunctival Grafts, Lid Surgery, and Neurotization. Eye Contact Lens. 2021;47(3):149–153. doi:10.1097/ICL.0000000000000753

25. Park JK, Charlson ES, Leyngold I, Kossler AL. Corneal Neurotization: A Review of Pathophysiology and Outcomes. Ophthal Plast Reconstr Surg. 2020;36(5):431–437. doi:10.1097/IOP.0000000000001583

26. Solyman O, Elhusseiny AM, Ali SF, Allen R. A Review of Pediatric Corneal Neurotization. Int Ophthalmol Clin. 2022;62(1):83–94. doi:10.1097/IIO.0000000000000403

27. Sheha H, Tighe S, Hashem O, Hayashida Y. Update On Cenegermin Eye Drops In The Treatment Of Neurotrophic Keratitis. Clin Ophthalmol. 2019;Volume 13:1973–1980. doi:10.2147/OPTH.S185184

28. Hays RD, Bjorner JB, Revicki DA, Spritzer KL, Cella D. Development of physical and mental health summary scores from the patient-reported outcomes measurement information system (PROMIS) global items. Qual Life Res Int J Qual Life Asp Treat Care Rehabil. 2009;18(7):873–880. doi:10.1007/s11136-009-9496-9

